# SARS-Cov-2 proliferation: an analytical aggregate-level model

**DOI:** 10.1101/2020.08.20.20178301

**Authors:** Thomas Pitschel

## Abstract

An intuitive mathematical model describing the virus proliferation is presented and its parameters estimated from time series of observed reported CoViD-19 cases in Germany. The model replicates the main essential characteristics of the proliferation in a stylized form, and thus can support the systematic reasoning about interventional measures (or their lifting) that were discussed during summer and which currently become relevant again in some countries. The model differs in form from elementary SIR models, but is contained in the general Kermack-McKendrick (1927) model. It is maintained that (compared to elementary SIR models) the model is more faithfully representing real proliferation at the instantaneous level, leading to overall more plausible association of model parameters to physical transmission and recovery parameters. The main policy-oriented results are that (1) mitigation measures imposed in March 2020 in Germany were absolutely necessary to avoid health care resource exhaustion, (2) fast response is key to containment in case of renewed outbreaks. Two model generalizations aiming to better represent the true infectiousness profile and aiming to incorporate recurring susceptibility are stated and numerical results for the latter are presented.

## 1 Introduction

Construction of the model has been motivated in course of the analysis of intensive-care capacity expenditure to be expected from sector-specific lifting of restrictions. This former analysis used a budget-oriented argument to arrive at an *indicative estimate* of the resource expenditure, but did not analyze dynamics. (Concretely it assumed a constant rate of new infections.) A reasonable question to be posed is: If a sector was allowed to reopen, what would the trajectory of infections actually look like, when surely it is not a linear increase? Further, can parameters of the local transmission behaviour be derived from the aggregate observed numbers?

In the present text, a model capturing the dynamics of the number of infections is developed towards answering these questions. It deliberately contains only few parameters and is in fact not designed to a specific stage of the virus proliferation. Though models for tracing the trajectory of infectious diseases exist, for example the intuitive SIR model [Ken56] which is formulated as a system of scalar differential equations, it is maintained here that physically more realistic descriptions are possible, which, moreover, lead to increased accuracy of estimated parameters.

Section 2 and 3 describe the model and underlying reasoning in detail, state scaling properties useful in fitting the model and explore qualitative features of resulting trajectories. In section 4, the model is fitted to the evolution of the epidemic in Germany as observed via reported CoViD-19 cases until mid-May 2020. Section 5 states policy insights. Finally, two appendices describe model extensions, of which the first is relevant to taking into account asymptomatic cases.

## 2 The model

We assume a homogeneous set of individuals which act as unwitting agents in the proliferation. We assume that infection spreads probabilistically from the infected (and still contagious) individuals to any other individual of the set, wherein we assume that each individual is connected randomly to others, but such that all individuals approximately have an equal number of neighbours (=: A1). (In graph-theoretic terminology, the graph of contacts between individuals is a random undirected graph where each node has about the same edge degree *d*.) No other assumptions are imposed on the global topology of interconnections. Individuals who were once infected cannot be infected again (=: A2). Finally, an assumption here made is that contagiousness lasts only for a duration *t_c_*, i.e. an infected individual is contagious for the period [0,*t_c_*] after its infection and then not at all afterwards (A3). This simplified characteristic is motivated by results on infectiousness found in epidemiological and clinical investigations: In [HLWea20], infection incidence data of the Wuhan area is examined and combined with clinical data to derive an infectiousness profile which has most of its weight located at about 7 consecutive days around the symptom onset^1^. [WCG+20] recorded viral RNA load data in sputum, throat swab and stool and report of nine patients viral peak loads of 2.35 · 10^9^ copies per *ml* sputum, declining rapidly starting from the first day of presentation in almost all patients, decreasing to 10^5^ copies per *ml* within about 10 to 16 days after symptom onset. (A level of below 10^5^ copies per *ml* sputum, combined with no symptoms and past day 10, has been regarded as warranting discharge of the patient from clinical care with ensuing home isolation.) [TTL+20] (Fig 2) report viral load in posterior oropharyngeal saliva samples decreasing monotonously to below 10^4^ copies per *ml* in day 21 after symptom onset, for the majority of 20 nonintubated patients (out of *n* = 23). Changes in population size due to non-disease effects will be ignored, instead *N* will be considered constant; similarly the changes in proliferation characteristic due to disease-related reduction of the population will be deemed negligible. Assumptions A1 to A3 will be “baseline” assumptions throughout the text and substantial deviations from them will be discussed in the appendix only.

**Figure 1:**
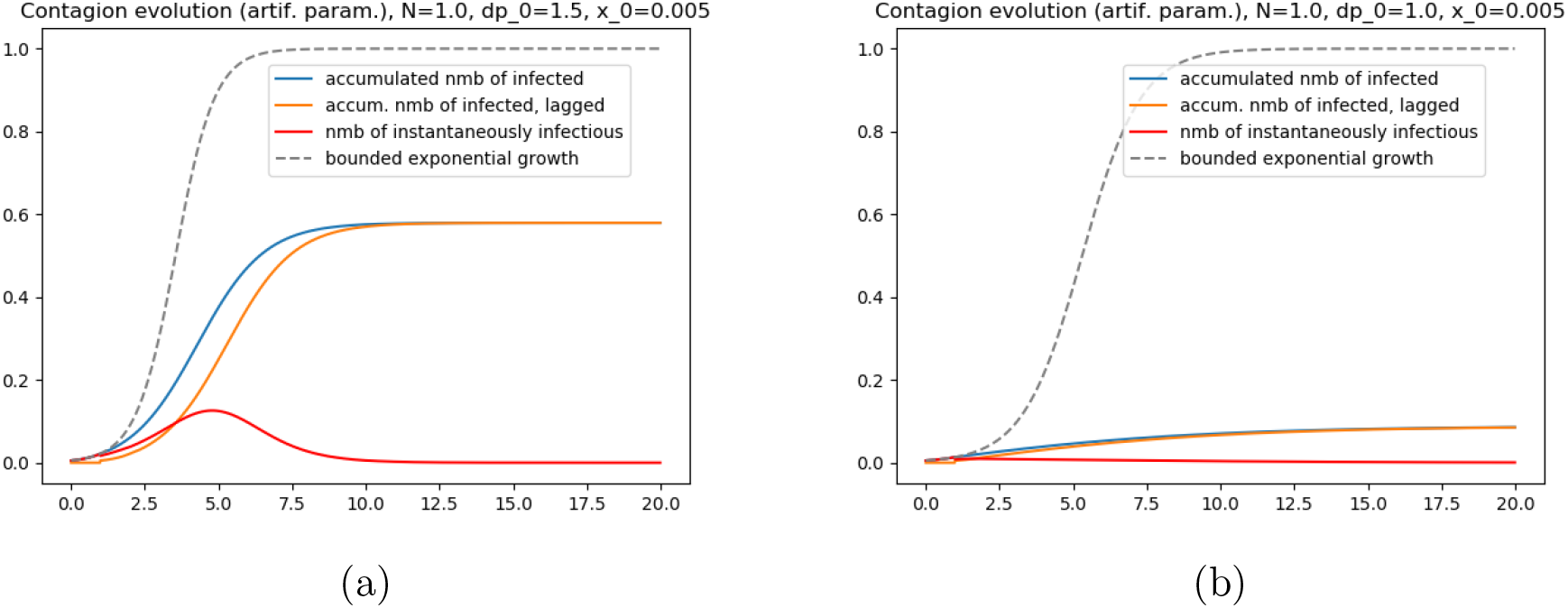
The solution obtained for equation (2) with *x*_0_ =0.005 and using (a) *d · p* =1.5 or (b) *d · p* =1.0. The blue solid line shows the accumulated number of infections, and the orange solid line the same curve but delayed by *t*c = 1 (which denotes the number of individuals once having been infected but not anymore being contagious). The red line shows the number of instantaneously infectious. Crucially, the accumulated number of infected does not increase to fully exhaust *N*. For comparison, the grey line shows the accumulated infections if the infectiousness did not cease after *t*c.

**Figure 2:**
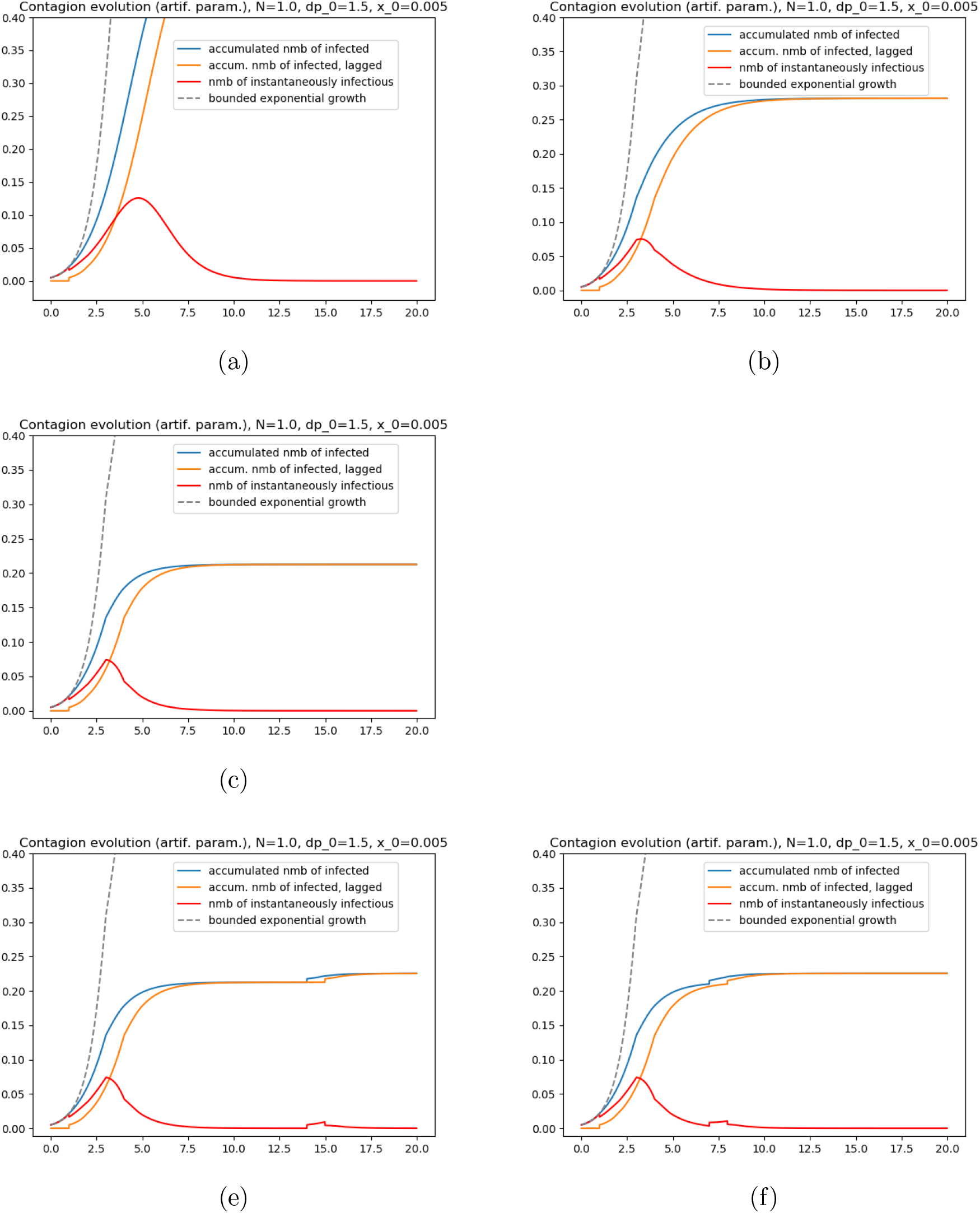
(a) Detail view of the solution in Figure 1a. (b) Solution obtained when using *d · p* =1.5 for *t* ∈ [0, 3), then *d · p* =1.0 for *t* ≥ 3. At *t* = 4 there is a “bend” in the graph of the increment of the number of infections (red line). The bend naturally occurs after duration *t*c after the switch of the parameter values was made. (c) Solution as in 2b but using *d · p* =0.8 for *t* ≥ 3. (e) Solution obtained when simulating a second (overlaid) outbreak event (of same strength as the initial one, i.e. Δ*x* = *x*_0_), at *t*_2_ = 14.0. (f) Solution with the second outbreak occuring at *t*_2_ =7.0. Noteworthy is (in both cases) that even though the same number of exogenously infected was used as initially, the contagion effect is much smaller. This is because at that stage and with this parameter setting, already about one fifth the population had been infected (thus was immune in this model).

We aim for a numerical formulation of the aggregate evolution in which the randomness is averaged out. For this, let *N* be the number of agents, and let at *t* = 0 the number of infected agents *x*(*t*) be given as *x*_0_ *<N*. Before *t* = 0, the number of infected agents shall be zero. To develop the model incrementally, lets momentarily assume that all infected agents are contagious infinitely long. In a unit time interval, all infected agents are deemed to infect each of respectively *d* other neighbours – stochastically independently – with probability *p*. The expected total number of virus receivers, per unit time interval, then is *x*(*t*) · *d · p*. But not all receivers get infected because some are already infected. The share of non-infected receivers among all agents is (1 − *x*(*t*)*/N*); therefore the expected number of new infections in unit time is *x*(*t*) · *d · p ·* (1 − *x*(*t*)*/N*).

Approximating the model evolution as continuous process even at small time intervals (reasonable given the size of the numbers involved), one concludes, under assumption of infinitely enduring contagiousness, that *x*(*t*) follows

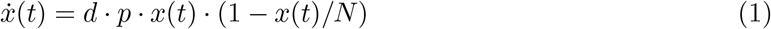

for *t* ≥ 0, with *x*((−∞, 0))=0 and *x*(0) = *x*_0_. Obviously the function *x*(*t*) is non-decreasing.

For incorporating the finite duration contagiousness, one determines the number of contagious individuals as the difference of the accumulated number of infected at time *t* minus the accumulated number of infected prevailing at the earlier time *t*−*t_c_*, because that share of agents had the infection already for at least duration *t_c_*, and will cease to be infectious at *t*. Consequently, the expected total number of virus receivers is refined towards (*x*(*t*) − *x*(*t* − *t_c_*)) · *d · p*. The model with the finite duration contagiousness thus reads, in expectation,

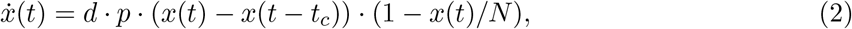

with initial conditions as before. Both differential equations respectively have a unique solution. ^2^

### 2.1 Relation to existing models

The here presented model is not representable by the elementary SIR models that involve only instantaneous evaluations of the state variables on the right-hand side of the differential equation, as e.g. equation (2) in [Ken56] (see [ZML+20] for a current example of its usage). It is therefore also necessarily different for example from [MB20]. The reason for this is a restriction imposed by such formulations, namely that the individual’s transition from infection to recovery is modelled using a *rate* of transition *proportional* to the number of infected individuals, which corresponds to a stochastic recovery occurrence and yields an exponential decay characteristic on average. It is known however that, in reality, the SARS-CoV-2 shows a rather deterministic disease progression with regards to infectiousness in time, leading to end of the infectiousness after about two to three weeks after begin of infection, based on cell culture (see earlier citations). This clinically supported characteristic is properly represented in equation (2), but not in elementary SIR models.

On the other hand the here presented model *is* conceptually contained in the original (i.e. general) compartmental model of Kermack and McKendrick [KM27] (which involves a formulation using integrals; see comments in [Bra17] also), for example by setting there *ψ* = 0. ^3^ This holds also for the refinement given in section B.

The advantage of the here given formulation is that it allows for a mathematically relatively simple description while still fully allowing accomodation of the infectiousness characteristic in generalized form. This simplicity gives some room to incorporate other, hithertho unconsidered, effects into the model and still retain a model complexity which is amenable to simulation for parameter identification.

## 3 Analysis of the model and exploratory simulation

For later simulation, it is helpful to make use of the scale invariances inherent in the above differential equations. If one denotes the equation (2) parametrized with *d·p* and *t_c_* and *N* and initial value *x*_0_ as “ODE(*dp, t_c_, N, x*_0_)”, then we have the following fact: If *t* ↦ *x*(*t*) is a solution to ODE(*dp, t_c_, N, x*_0_), then *t* ↦ *x*(*at*) is a solution to ODE(*a · dp, t_c_/a, N, x*_0_) for any *a>* 0. This means we can restrict analysis for example to *t_c_* = 1 and vary only *d · p* and *x*_0_.

The other scale invariance is described by “*x*(·) solution of ODE(*dp, t_c_, N, x*_0_) then *ν · x*(·) is solution of ODE(*dp, t_c_,ν · N, ν · x*_0_)”, where *ν ≠* 0.

Instead of a further analytical proceeding, the above equation’s evolution was examined via computer simulation, for various parameter choices *d · p* and initial values. The purpose is first to explore the general (i.e. not real-data matched) behaviour of equation (2) (next subsection), then to fit the parameters to observed real data (section 4). Throughout it was used *N* =1.0, *t_c_* = 1 and a (forward Euler) discretization step size of 0.01 (corresponding to `resolution=100` in code).

### 3.1 General model behaviour

The below discusses general features of the model and its behaviour under parameter variations. This is for demonstration only, and arguments on the proliferation phenomena should be taken as schematic. (Whether the phenomena occur in the real parametrization is to be discussed in section 4.)

Fig 1a shows the evolution behaviour for some arbitrary but temporally constant parameter set. The most striking feature at this graph is that the number of infections asymptotically does *not* reach the total number *N* of agents. Rather, the limit is a value *x*(∞) *<N* which depends on the *d · p* and the initial value. For comparison, the evolution of the number of infections as would arise when observing eqn. (1) [with same *d · p* parameter] is depicted as grey dashed line; in it, the *x*(*t*) converges to *N* independent of the choice of *d · p*. (In subsequent text, this will be referred to as “bounded exponential growth”.) The reason for including this curve here and in following graphs is that it can give a hint on trajectories of future viruses that may have a much more extended infectiousness interval. In fact, this curve would result if infected individuals remained infinitely long infectious and were not quarantined.

#### Dependence on parameters

In simulations, the dependence of the limit *x*(∞) on *d · p* appeared to be generally over-proportional (see Fig 1b). This is well-known behaviour also in the instantaneous-state models. On the other hand, the dependence of the limit on *x*_0_ was linear or sub-linear. In instantaneous-state models, the limit does not depend on the size of the initiating jump of *x* at *t* = 0. (check)

#### Nearly linear growth of infections for a substantial time period can be represented

Prolonged linear growth of infections, after initial exponential growth, is exhibited for suitable parameter choices of *d · p* and *x*_0_. See section 4, Fig 4.

**Figure 3:**
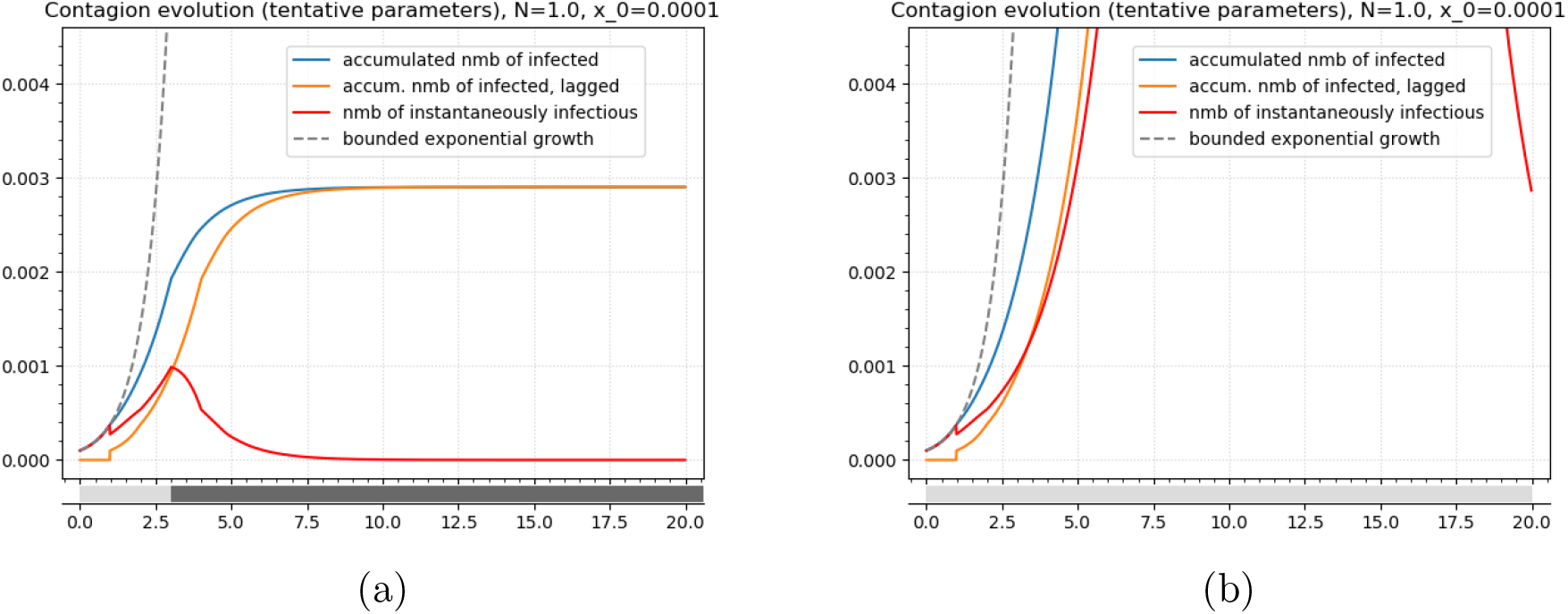
Tracing reported CoViD-19 infections in Germany. As in Figures 1 and 2, the ordinate values denote the share of total population. The time scale is chosen such that [0, 1] corresponds to two weeks. (a) As Fig 2, but using initial value *x*_0_ =0.0001 and initial *d · p* =1.34. For *t* ≥ 3 the *d · p* is 0.65. The simulation yields *x*(∞) ≈ 0.00290. (b) Shows the evolution as would occur if the parameter switch at *t* =3.0 was omitted. Then *x*(∞) ≈ 0.44520.

**Figure 4:**
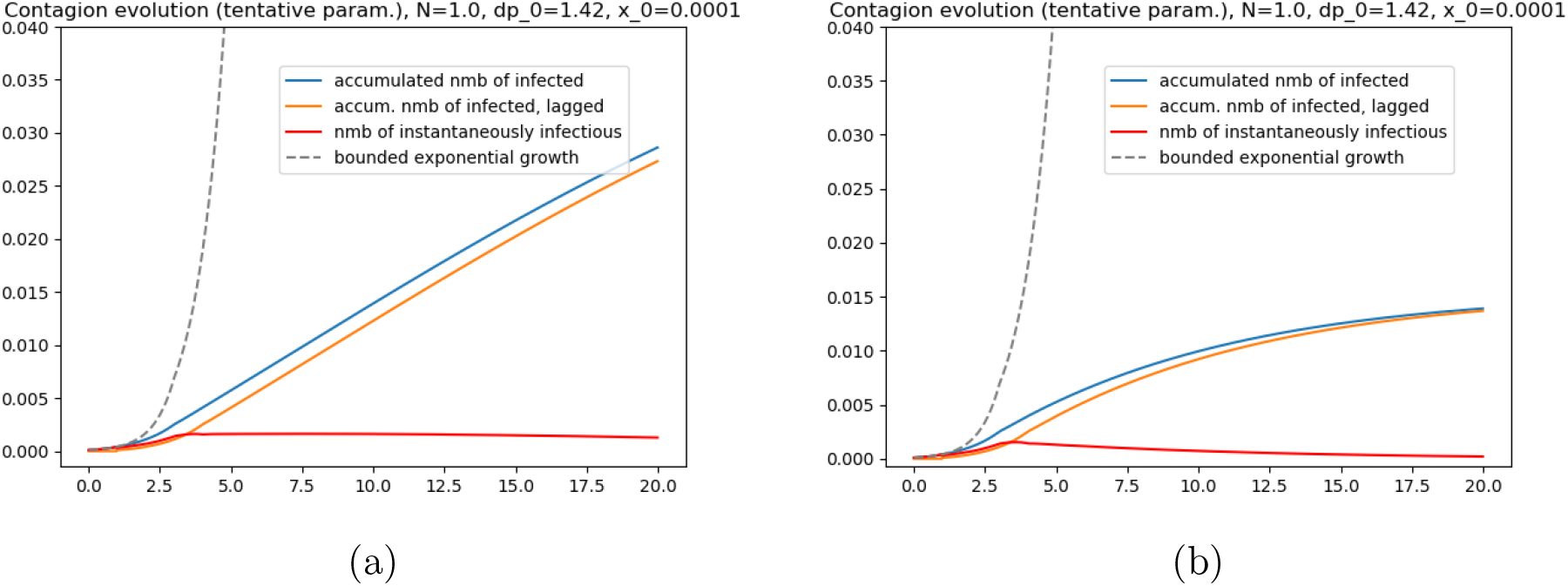
(a) Solution for a parameter choice such that *x*(*t*) shows a pronounced phase of nearly linear growth: it was *d · p* =1.42 initially, then *d · p* =1.02 from *t* =3.0 onwards. Not depicted: Also with this parameter choice the *x*(*t*) eventually converges: the linear growth phase extends until about *t* = 25.0, and finally it is *x*(∞) ≈ 0.04948. (b) Solution obtained when using the same initial conditions, but switching to *d · p* =0.96 at *t* =3.0.

#### Behaviour upon later occurrence of a second virus source

An interesting question is how the system behaves if a second outbreak is occuring at a later time when the number of infections *x*(*t*) already has grown substantially. Fig 2e and 2f show the results for two different choices for the moment *t*_2_ of the second initiation. It is noteworthy first that a second outbreak does not lead to substantially more infections. Second, the *x*(∞) depends on this *t*_2_ only minorly. The values are *x*(∞)=0.225490 for *t*_2_ = 14.0, *x*(∞)=0.225491 for *t*_2_ =7.0, *x*(∞)=0.23732 for *t*_2_ =2.0, and (for reference) *x*(∞)=0.3114 for *t*_2_ =0.0.

## 4 Modelling real infectedness trajectories

We use here the number of reported CoViD-19 cases (as aggregated by the Robert-Koch-Institut [Rob], available also in [ECD20]) as a proxy for the number of infections in Germany.^4^ We fit parameters for the interval until beginning of May 2020, assuming that the evolution proceeded within two different parameter regimes: first a *d · p* corresponding to no restrictions, then a *d · p* corresponding to the restrictions posed by contact disencouragement and store closure. (The observational interval used for parameter estimation does cover only a few days of the time of obligatory indoor face mask wearing.) We can derive parameters and based on them predict the trajectory of infections way forward. Because of the simplicity of the examined model, there is the risk of a high model error existing. Therefore, at the present state of this text, such estimation can only serve to determine reasonable bounds on the parameters of the model, rather than to give a reliable forecast of expect number of eventual infections.^5^

### Parameter fitting

Fitting of parameters is here conducted manually, focussing on moments in the time series that are indicative of parameter changes. At the beginning of April 2020, the number of weekly new CoViD19 cases stood at about 40000 in Germany. If we regard the modelling time unit to correspond to a real duration of 2 weeks (implying that each individual newly infected is non-contagious two weeks after and onwards), then we have a new-infections rate of 80000 individuals per such time unit which corresponds to an increment of approximately Δ*x* =0.001 per unit time after normalizing to *N* =1.0. Identifying the moment which was one week after the initial wider lock-down in Germany (i.e. around 29th March) as moment *t* = 3 in the modelling, parameters consequently need to be fitted such that *ẋ*(3.0+) = 0.001 (red line). (The *t* =3.0 also implies that the model assumes around 6 weeks of initial evolution under a low-restrictions scenario, which matches the timeline of the outbreak in Germany approximately.) Fig 3a shows the trajectory of the system evolution using initially *d · p* =1.34 and switching to *d · p* =0.65 afterwards. Fig 3b shows the evolution if no parameter switch (i.e. no intervention) had happened at *t* =3.0.

Note: The matching is overly simplified for the interval *t* ∈ [0, 3.0], leading to an overestimated *x*(*t*), since for example *x*(3.0) ≈ 0.0020 –corresponding to 160000 individuals–, while the actually reported number was around 52550. In reality, the *d · p* will have varied in that interval, with the natural (but not proven) assumption being that towards the end of [0, 3.0], the parameter was lower than its temporal average.

### Intensive care capacity

In order to derive a maximum allowable number of new infections for evaluating strategy choices, the expected implied number of cases requiring intensive care (in particular artificial ventilation) must be limited versus the available capacity of intensive-care units (”ICU”), with the non-Covid-19 patients’ demand taken into account. Based on data from the DIVI IntensivRegister [DIV20], as of 15th Sept. 2020 the number of existing ICUs in Germany is 30645 (from 1283 reporting sites), 21836 of these are occupied and in turn of these 236 are occupied by Covid-19 patients. (The occupancy by non-Covid-19 patients apparently increased by about 6000 during the months April to September 2020 but by now has levelled.) As an approximation suitable for this text, it is reasonable to use 9045 as the available ICU capacity for Covid-19 cases. Further, the ratio of ICU-requiring Covid19 cases to all reported new infections is deemed to be about 5% here, using a prudent estimate computed from the DIVI IntensivRegister data and new infections data. It is worth to note that this ratio depends on the age distribution among the new Covid-19 cases, which has shifted to younger people in recent months. Combining the two numbers and an estimated average Covid-19 patient occupancy duration of two weeks yields that the maximum allowable number of weekly new infections is 9045*/*5%*/*2 = 90450, or 180900 per two weeks, which corresponds to ordinate value ≈ 0.00226 in the graphs of Fig 3. ^6^

### Concerning evolution upon potential second outbreak

It is necessary to remark that the conclusion drawn in connection with Fig 2e and 2f – i.e. that a second outbreak of similar magnitude as initially would not effect a substantial increase in the accumulated number of infected individuals – cannot be affirmed for the current scenario (in Germany and elsewhere), since that number is rather about 0.25% to 0.5% of total population currently, rather than the 1*/*5 prevailing in the demo scenario in Fig 2e and 2f at the onset of the second outbreak.

## 5 Policy insights

The challenge with lockdown measures for the current corona virus is the following: When imposing them, they will show effect only if the basic reproduction number is pushed below 1 sufficiently enough. Then, after the number of infected individuals has eventually dwindled, a lift of the lockdown is tempting - however even a slight increase of *R*_0_ above one opens the way to renewed catastrophic infections increase. One therefore has a binary evolution characteristic; to control *R*_0_ by policy such that a steady stream of just managable new infections is maintained is daunting, and likely impossible (in practice) if a policy requiring a constant set of restrictions is targetted. The natural answer, at least from a theoretical point of view, is to consider phases of lifted restrictions interleft with repeated adaptively switched phases of more stringent restrictions or more stringent enforcement of existing restrictions. The need for such strategy is not in principle altered by the local aspect of transmission, except that switched lockdowns only need to be local and thus do not affect the whole population.

Further, the graphs suggest that a future virus having infectiousness lasting much longer than the about 2 to 3 weeks for SARS-Cov-2 and also being as highly infectious would pose serious challenges (if the effective infectiousness period also increases), because of resource exhaustion in the mid-stages of the pandemic.

## 6 Conclusion

In this study a partly novel model for virus proliferation dynamics was developed and with it the SARS-Cov-2 outbreak in Germany retraced on an aggregate level, using CoViD-19 case count data by the Robert-Koch Institute in Berlin. Elementary properties of the model were identified. Predictions by the model for different levels of mitigation measures were hinted at or stated in approximate manner, and put into context of available health care resources in Germany.

Future policy oriented work would need to address better understanding of fine-grained and adaptively activated mitigation measures, for which a spatial model should be favoured over purely aggregate models as the present one. Further, for purpose of improving parameter and state estimates, the issue of underreporting (i.e. #actual *>* #reported cases) must be taken into account appropriately. Ideally, one can develop an estimate for the factor of underreporting from more exact spatial analyses.

On the mathematical side, a more rigorous formulation of the instantaneous proliferation dynamics is desirable, which allows to link parameters of the aggregate model to well-defined elementary parameters and results in more systematic parameter estimation. The ultimate goal is to be able to estimate more local structure from the observed time series.

## Data Availability

Raw input data used have been obtained from Robert-Koch-Institut, Berlin. (see link)
An alternative source for the same data is the European Centre for Disease Prevention and Control.

https://www.rki.de/DE/Content/InfAZ/N/Neuartiges_Coronavirus/Fallzahlen.html

https://data.europa.eu/euodp/en/data/dataset/covid-19-coronavirus-data

## A Data series on daily newly reported CoViD-19 cases (Germany)

**Figure 5:**
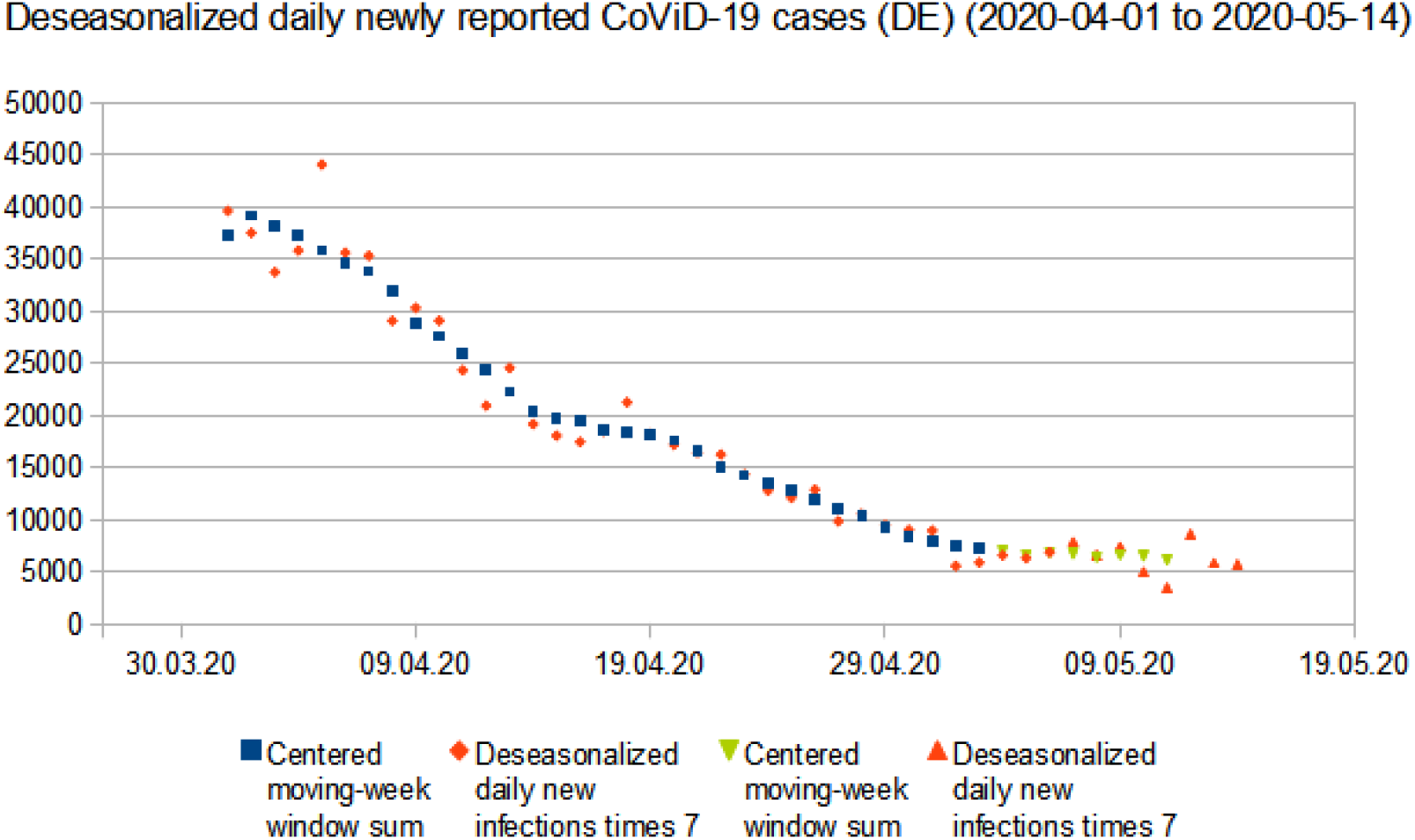
A “smoothed” derivate of numbers of daily newly reported CoViD-19 cases in Germany published by [Rob], see [ECD20] also. The blue squares and green triangles series show (for comparison) the sum of daily new cases over a moving 7-day window. Orange diamonds and triangles show daily new cases after scaled with a weekday-specific weight factor to remove the weekly pattern seen in the original data. The weight factors were estimated from data corresponding to the squares and diamonds series, i.e. from the interval from 1st April until 6th May 2020. Germany imposed face-mask wearing in stores starting from 27th April and allowed certain (moderate) shop reopening starting from 4th May 2020. The “bend” at around 14th April is remarkable because no changes in measures were effected at that time or within the preceding one week.

## B Refinement of the infectiousness mechanism

So far, a crude specification of the infectiousness has been used, putting focus on the typical *effective* infectiousness interval of about two weeks. Further aspects in the virus transmission which could be accounted for in a refinement are the transmission from longer lived remnants of the virus in otherwise recovered individuals^7^, or – tractable along a similar line – the transmission from asymptomatic cases.^8^

Examining the transmission from viral remnants first, imagine that individuals infected at time *t*_0_ remain contagious until *t*_0_ + *t_c_*_2_ with reduced probability, additionally to the previously used interval [0,*t_c_*]. Concretely, let *p*_2_ be the probability that an individual which has been infected for a duration exceeding *t_c_* but not exceeding *t_c_*_2_, will transmit the virus in a unit time step. With 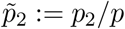 the adjusted model equation then reads

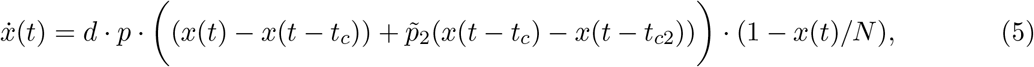

since those individuals must be added to the instantaneous reservoir from which infections are generated. The equation is better written as

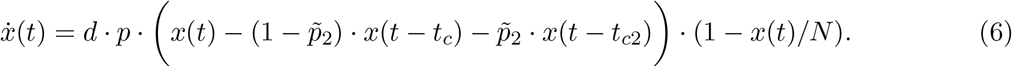

If we denote by *i*(*t*) the infectiousness profile, which shall describe the relative infectiousness of an infected individual^9^ at time increment +*t* after the infection moment (relative to infectiousness at *t* = 0), then the above used specification for SARS-Cov-2 is expressed as

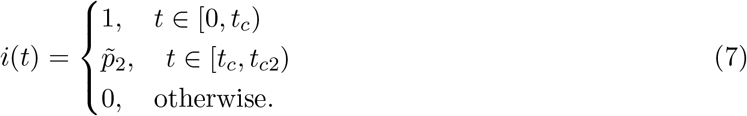

Its derivative is (with Dirac notation) 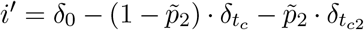. One therefore finds that the model equation (6) in fact is generally best written as

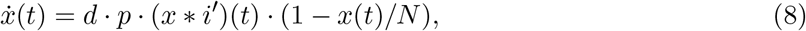

where “∗” denotes the function convolution. A Dirac notation-free representation derives from (*x* ∗ *i′*)(*t*)= ∫_ℝ_ *x*(*t* − *s*)*i′*(*s*)d*s* = ∫_ℝ_ *x*(*t* − *s*)d*i*(*s*). Here the last integral signifies the well-known Stieltjes integral.

Note: The infection from contaminated surfaces of objects can be represented in the same framework. This is because initially and during the evolution of the spread, viruses are on surfaces mostly there where infected individuals previously had been.

## C Incorporating renewed susceptibility

In the literature, it was mentioned that a substantial fraction of previously deemed cured patients were testing positive again on viral RNA a short period after discharge from SARS-Cov-2 related treatment [HDG20] [YLea20]. In different context, the permanence of antibodies in the blood after recovery has been questioned and found to last only two to three months [LTea20]. Relevant from the epidemiological viewpoint is whether “re-positive” cases are infectious, and according to the evidence this seems only to be true upon reinfection after decrease of antibody levels. In this section, the model is to be extended to cover this reinfection mechanism, i.e. focus here is on the route via resusceptibility after loss of antibodies.^10^

When transition from the recovered state to the susceptible state is to be accounted for, the modelling using a univariate state variable cannot represent the new infections by a change of *x*(*t*), if that is to represent the total number of at time *t* ever infected people. Introduction of a second state variable *r*(*t*), which shall denote the number of renewed susceptible individuals (multiple times counted if multiple such “resets” happen on one individual over time), will allow to extend the original model to make it represent the new mechanism. With *x*(*t*) denoting the number of infected individuals (again multiply counted if necessary), the *x*(*t*) − *r*(*t*) will denote the number of individuals (at *t*) being either “infected” or “recovered and not newly susceptible”. I.e., this is the share of the whole population in which virus transmission does not effect an infection. (=: “instantaneously non-susceptibles”) Therefore the updated equation for evolution of *x*(*t*) reads

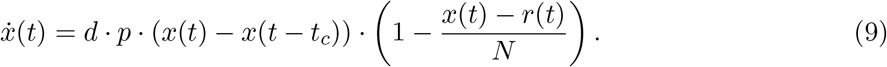

By definition, any individual counted in *r*(*t*) must have been counted in *x*(*t*) before, so *x*(*t*) ≥ *r*(*t*) for all *t* must be ensured in the modelling of the evolution of *r*(*t*). Beyond this minimal requirement, it is assumed here a mechanism that demands: (A1’) the transition to renewed susceptibility does not begin before a duration *t_r_* has elapsed starting at the infection moment, and (A2’) the transition then proceeds with a constant rate *α_r_* per time unit.

Under these assumptions, an appropriate evolution equation for *r*(*t*) is given by^11^

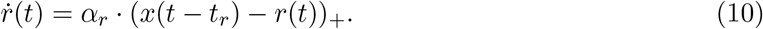

The subscript “+” denotes the “positive part” of the argument. By construction, the share of potentially reverting individuals that does actually revert to susceptibility in one time unit is designed to be 1 − exp(−*α_r_*). (Example: *α_r_* =0.2/week means about 18.1% of potentially reverting do actually revert to susceptibility in one week.) For consistency, it must be ensured *t_r_ >t_c_*, or, when the adjusted generalized model with eqn. (8) were to be used, *t_r_ >* max{*s* ∈ ℝ*,i*(*s*) *>* 0}. The “safe period”, i.e. from end of infectiousness to start of possible reversion to susceptibility, lasts for a duration *t_r_* − *t_c_*.

With this model given by equations (9) and (10), employing again the time unit of two weeks, and using the values *t_r_* =4.0 and *α_r_* =0.2 (corresponding to six weeks of safe period rate of reversion of 0.2 per two weeks), one obtains numerical results as in Fig 6. The value of *d · p* was: 1.34 until *t* = 3, then 0.77 until *t* = 7 (representing the major lockdown period starting near end of March 2020 in Germany). The whole abscissa corresponds to somewhat less than four years. The level of maximum intensive care capacity in Germany (posing a limit to the red trajectory) again is represented by a horizontal line in the upper half of the graphs, implied from a maximum of 12500 to 25000 ICU cases per week. As before, spatial pecularities of the transmission or annual seasonal effects are not represented in the result. Possible future vaccination campaigns, which are also not represented here, will effect a reduction of parameter *d*.

Overall, even a qualitative difference in the evolution is visible in particular for the (here focussed on) case of *d · p* ≈ 1.0: If resusceptibility is possible, then transmission coefficient values slightly above 1.0 still lead to an increase of infectious cases (red line) close to or even above the hospital maximum resource level. On the other hand, only a slight decrease of the transmission coefficient (e.g. by slightly improved face mask discipline) will – according to the model – take the evolution into safe regions. Side note: The outcome depended only little on the parameter *t_r_* varied in [4.0, 8.0] (reflecting a hypothesized “safe period” of six to 14 weeks).

**Figure 6:**
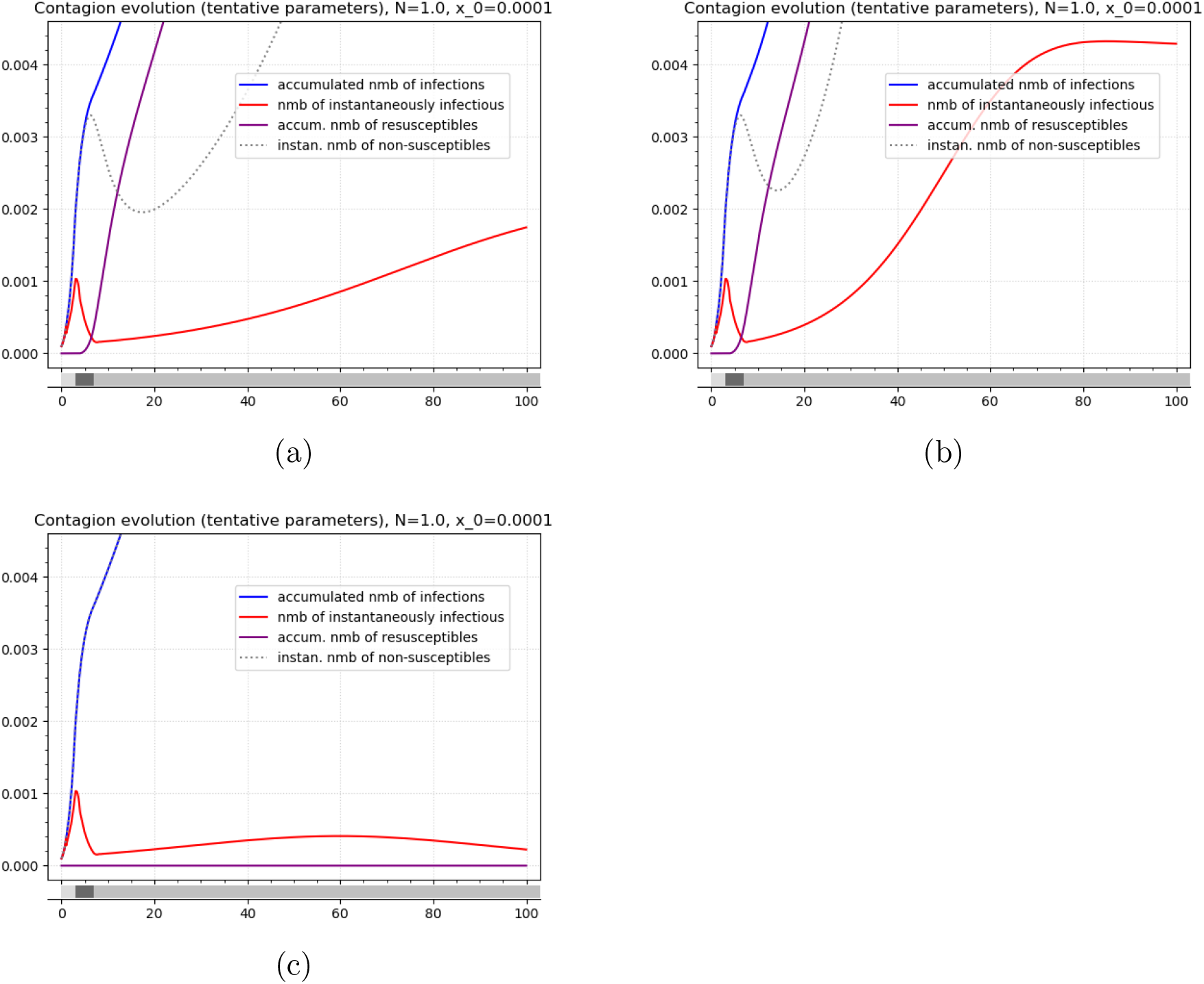
Proliferation evolution with renewed susceptibility possible. Abscissa: time with [0, 1] corresponding to two weeks; ordinate: share of population. The differently shaded regions at the bottom indicate phases of different *d · p* values. It was chosen *d · p* =1.34 until *t* = 3, then = 0.77 until *t* = 7. For *t* ≥ 7, it was *d · p* =1.02 in Fig a and *d · p* =1.04 in Fig b. Both figures assume parameters *t_c_* =1.0, *t_r_* =4.0, and *α_r_* =0.2. For comparison, Fig c shows the otherwise equal scenario as in Fig a, but without resusceptibility (i.e. *α_r_* = 0). Not depicted: In Fig b, the number of instantaneously non-susceptibles converges to about 3.9% of the population as *t* → 100.0; the number of ever infected at that time is about one fourth of the population.

## Footnotes

1 Caution in the usage of numbers from pure incidence analysis is required: As consequence of the way the raw data is obtained in [HLWea20], only infectiousness *around the moment of symptom onset* is in fact fully observed. This is because earlier transmission are likely usually not properly associated to the real primary case because the primary case does not show symptoms yet. Later transmissions are simply inhibited because the primary case is put into quarantine. An epidemiological analysis of incidence data alone therefore necessarily is insufficient to determine “pure” infectiousness. To emphasize the distinction between “pure”/”medical” infectiousness and infectiousness after taking into account the population’s socio-characteristics (household structures, current mitigation policies), it is worthwhile to call the density of the latter an “infection incidence profile”.

2 Instead of considering only a temporally finite and uniform infectiousness, more detail can be incorporated into the differential equation using a convolution term, as shown in appendix B.

3 With *ψ* = 0, have *C_θ_* = 0. Consequently *y_t_* = *N* − *x_t_* for all *t*. It follows 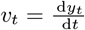 and 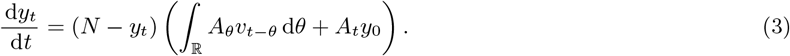 With 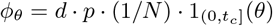 obtain 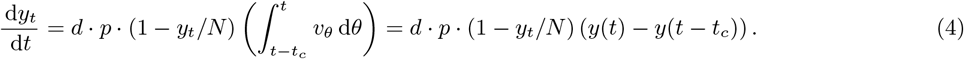

4 The issue of distinguishing between actual and reported infections is not taken to full length in this text. An ad-hoc approach is to scale the observed reported numbers to an “actual infections” estimate using a hypothetical factor, and then perform parameter estimation to match this hypothetical “actual infections” time series.

5 Two relevant aspects not represented in the model forecasts are the annual seasonality in the transmission coefficient and parameter changes due to changes in the *spatial* distribution of infection hotspots. Moreover, vaccination campaigns reduce *d* by an amount corresponding to the fraction of the population being vaccinated.

6 In a previous version of this text, the number of all ICU beds in Germany was approximated as 25000 and erroneously used as full capacity available for Covid-19 patients. Besides, the ratio of ICU-requiring Covid-19 cases to all reported new infections was assumed to be 1:7, based on data derived from the very initial stage of the proliferation in Germany, but meanwhile has decreased. Correcting for both issues happens to have a compensating effect such that the original statement remains nearly intact.

7 But see comments in next section also. Virus remants neither in self-infection nor in transmission seem to play a major role.

8 In asymptomatic cases, the effective infectiousness interval extends over the whole period of virus activity in the subject.

9 On the contrary, the profile in shown in Fig 1.c of [HLWea20] is best called an “infection incidence profile”, as explained on page 2.

10 The mechanism of *reactivation* of the virus in former patients must be represented in markedly different form, since individuals affected would pass directly from the “cured” to the “infected” state. For this, a further summand must be included on the right-hand side of the evolution equation for *x*(*t*), i.e. in eqn. (2) or (9). Note however that this refers only to the case of reattained infectiousness, which according to evidence [YLea20] is not the typical course. On the contrary, the mere detection of viral RNA (i.e. positive RT-PCR test) after cure typically indicates a shedding of inactive viral RNA, e.g. fragments.

11 A (so far minor) shortcoming of this modelling approach is that different probabilities of infection risk in first-time virus receivers and repeat virus receivers cannot be represented.

